# Management of Antipsychotics in Primary Care: Insights from Healthcare Professionals and Policy Makers in the UK

**DOI:** 10.1101/2023.11.13.23298487

**Authors:** Alan A. Woodall, Aseel S. Abuzour, Samantha A. Wilson, Frances S. Mair, Iain Buchan, Sally B. Sheard, Paul Atkinson, Dan W. Joyce, Pyers Symon, Lauren E. Walker

## Abstract

**Introduction:** Antipsychotic medication is increasingly prescribed to patients with serious mental illness. Patients with serious mental illness often have cardiovascular and metabolic comorbidities, and antipsychotics independently increase the risk of cardiometabolic disease. Despite this, many patients prescribed antipsychotics are discharged to primary care without planned psychiatric review. We explore perceptions of healthcare professionals and managers/planners of policy regarding management of antipsychotics in primary care.

**Methods:** Qualitative study using semi-structured interviews with 11 general practitioners (GPs), 8 psychiatrists, and 11 managers/planners of policy in the United Kingdom. Interviews were studied using inductive thematic analysis.

**Results:** Respondents reported competency gaps that impaired ability to manage patients prescribed antipsychotic medications holistically, arising from inadequate postgraduate training and professional development. GPs lacked confidence to manage antipsychotic medications alone; psychiatrists lacked skills to address cardiometabolic risks and did not perceive this as their role. Communication barriers, lack of integrated care records, limited psychology provision, lowered expectation of patients with serious mental illness by professionals, and pressure to discharge from hospital resulted in patients in primary care becoming ‘trapped’ on antipsychotics, inhibiting opportunities to deprescribe. Organisational and contractual barriers between organisations exacerbate this risk, with socioeconomic deprivation and lack of access to non-pharmacological alternatives driving overprescribing. GPs and psychiatrists voiced professional fears of being blamed if an event causing harm occurred after stopping an antipsychotic, which inhibited deprescribing. A range of actions to overcome these barriers were suggested.

**Conclusions:** People prescribed antipsychotics experience a fragmented health system and suboptimal care. Many simple steps could be taken to improve quality of care for this population but inadequate availability of non-pharmacological alternatives and socioeconomic factors increasing mental distress need key policy changes to improve the current situation.

## INTRODUCTION

Psychiatrists usually prescribe antipsychotic medication (APM) to treat serious mental illness (SMI) such as schizophrenia and bipolar affective disorder. Despite treatment, patients with SMI die 10-20 years prematurely, mainly from cardiometabolic diseases and cancer [1–5]. In recent years, ‘atypical’ APMs are increasingly prescribed due to reports of fewer extrapyramidal effects [6], but they confer increased risk of obesity, type-2 diabetes mellitus, and cardiovascular disease [5, 7], and require ongoing clinical monitoring of cardiometabolic risk factors to ameliorate these risks, including body mass index, blood pressure, lipid and glucose levels and monitoring for QTc prolongation on electrocardiogram (ECG) [8–10]. While APM are licensed for SMI, increased prescribing for personality disorder [11], depression [12], anxiety [13] insomnia [14], dementia [15], learning disability [16] and autistic spectrum disorder [17] is occurring. Currently, in the UK, only quetiapine is licensed as a treatment for depression, and risperidone is licenced for short-term use to manage aggression in patients with moderate to severe Alzheimer’s dementia; all other uses are off-licence [18]. In some countries, such as Norway, APM are regularly initiated by GPs [19]. In the UK however, APM initiation is mainly undertaken by psychiatrists; traditionally, where the patient remains under secondary care (outpatient clinic or hospital care led by a psychiatrist), psychiatrists optimise APM, and GPs mainly monitor cardiometabolic risk [20, 21]. Unfortunately, many patients with SMI who attend psychiatric appointments are less likely to attend their GP for physical health follow-up [20, 21]. Many international health systems, mainly designed to manage acute rather than chronic illness, struggle to provide continuity of care to patients with chronic mental illness [22]. Patients taking APM deemed stable from a psychiatric perspective, or who do not have psychotic illnesses, are often discharged by psychiatrists to be managed solely by GPs, which blurs this traditional division of care and potentially leaves gaps in integrated care for these patients [23]. APM prescribing is increasing [6]; approximately 32% of patients with SMI who are often prescribed APM long-term do not have specialist psychiatric reviews [24]. This study explores the management of patients taking APM discharged to primary care. We explored the views of healthcare professionals (HCPs) who deliver this care and managers/planners or directors of policy (MDPs) of the health organisations where APM prescribing is occurring in the UK. The aim of this study was to identify potential barriers and facilitators of holistic care for people taking APM and views on the causes of increasing APM prescribing.

## METHODS

### Design and Setting

Qualitative study to explore views of professionals and policymakers involved in care of patients prescribed APM in primary care, by conducting semi-structured interviews with participants recruited from the UK. The ‘Consolidated criteria for reporting Qualitative Research’ guided study design and analysis of results [25].

### Participants and Recruitment

Participants were recruited from England, Scotland, and Wales between 1^st^ July 2022 and March 31^st^ 2023, and included GPs and psychiatrists; some held additional roles as service managers or policy directors. In addition to HCPs, NHS managers, government advisors and policy directors from stakeholders (e.g., mental health charities, Royal Colleges) were also invited. Purposive sampling identified potential participants, who were sent study information and an invite to participate. Up to three invites were sent; if no response was received, attempts to recruit ceased. Participants were briefed about the study with researchers and written consent obtained prior to interview. Participants could withdraw consent at any time, including after interview.

### Data Collection

Semi-structured interviews were undertaken between July 2022 and May 2023, and basic details of participant characteristics (professional role, whether clinicians had postgraduate experience of psychiatry and general practice, and duration of UK professional registration) were obtained. Interview schedules were developed and refined by the team (see supplemental file 1). Interviews were conducted by AW (clinician in general practice and psychiatry with qualitative research experience) and explored views on management of APM prescribing, barriers to holistic care and causes of increasing APM prevalence in the population. Interviews were conducted via Microsoft Teams. Audio data was transcribed verbatim and anonymised; each participant was assigned a code, then recordings deleted. Transcripts were not provided to participants for checking; no withdrawal of consent occurred after interview. Data collection and analysis were undertaken concurrently.

### Data Analysis

Interview transcripts were analysed using inductive thematic analysis, using Braun and Clarke’s methodology [26] to determine themes by constant comparison and ensuring data saturation was reached. QSR NVivo® Windows (Version 1.6) was used to organise, code, and analyse transcripts. Transcripts were repeatedly re-read to familiarise researchers with the data. This was followed by generation of initial codes, identifying themes and subthemes, and refining these to ensure final theme definitions reflected meaning within the data. The iterative categorisation of data into themes and subthemes, and interpretation of the data were undertaken by the coding team (AW, AA, SW, LW) to ensure analytical rigour and plausibility.

### Patient and Public Involvement

A public advisor (PS) provided input to protocol design, study progress, analysis, and interpretation of results.

### Ethics statement

The study received ethical approval from the Health Research Authority (IRAS: 311503) via NHS Research Ethics Committee-06 (22/WA/0173).

## RESULTS

Table 1 shows characteristics of participants; due to the national role of some MDPs, certain identifiable characteristics have been withheld to protect their anonymity. Of 46 subjects approached (20 GPs, 13 psychiatrists, 13 MDPs), 30 subjects (11 GPs, 8 psychiatrists, and 11 MDPs) consented to interviews, lasting between 21-58 minutes (mean 41 minutes).

**Table 1:** Characteristics of participants.

| Participant ID | Role | Sex | Postgraduate UK training experience in psychiatry and general practice? | Time on relevant UK professional register |
| --- | --- | --- | --- | --- |
| GP-01 | GP partner | Male | Yes | 21 years |
| GP-02 | Salaried GP | Male | Yes | 22 years |
| GP-03 | GP partner | Female | Yes | 16 years |
| GP-04 | Salaried GP | Male | Yes | 16 years |
| GP-05 | GP partner | Male | Yes | 21 years |
| GP-06 | GP partner | Male | No | 32 years |
| GP-07 | Salaried GP | Male | Yes | 8 years |
| GP-08 | GP partner | Female | Yes | 21 years |
| GP-09 | GP partner | Male | No | 11 years |
| GP-10 | GP partner | Female | Yes | 20 years |
| GP-11 | GP partner | Female | Yes | 21 years |
| PSY-01 | Consultant Psychiatrist | Male | No | 19 years |
| PSY-02 | Consultant Psychiatrist | Male | No | 25 years |
| PSY-03 | Associate Specialist in Psychiatry | Male | No | 30 years |
| PSY-04 | Consultant Nurse in Psychiatry (Responsible Clinician) | Male | No | 27 years |
| PSY-05 | Consultant Psychiatrist | Male | No | 21 years |
| PSY-06 | Consultant Psychiatrist | Female | No | 17 years |
| PSY-07 | Specialty doctor in Psychiatry | Male | No | 10 years |
| PSY-08 | Consultant Psychiatrist | Male | No | 22 years |
| MDP-01 | Former Educational advisor (psychiatry) to national postgraduate training committee and Royal College of Psychiatrists | Withheld | No (also a consultant psychiatrist) | Withheld |
| MDP-02 | Policy Director, mental health charity | Withheld | N/A | N/A |
| MDP-03 | Government advisor on mental health | Withheld | Withheld | Withheld |
| MDP-04 | Former executive officer of Royal College of General Practitioners | Withheld | Yes (also a GP) | Withheld |
| MDP-05 | Medical Director of Local Medical Committee for health region | Male | Yes (also a GP) | Withheld |
| MDP-06 | Director of Mental Health Services for provider organisation | Withheld | Withheld | Withheld |
| MDP-07 | Director of Primary Care for a provider organisation | Withheld | Withheld | Withheld |
| MDP-08 | Chief Pharmacist for a provider organisation | Withheld | Withheld | Withheld |
| MDP-09 | Medical Director for a provider organisation | Male | Yes (also a GP) | Withheld |
| MDP-10 | Representative of General Practitioners Committee (British Medical Association) | Withheld | Yes (also a GP) | Withheld |
| MDP-11 | Former executive officer of General Practitioner Committee of British Medical Association | Withheld | Yes (also a GP) | Withheld |

Five themes emerged that affected APM management in primary care: (1) Confidence of HCPs around holistic management of APM; (2) Service pressures ‘trapping’ patients on APM in primary care; (3) Communication barriers between primary and secondary care; (4) Lowered expectations of patients by HCPs; (5) Strategic factors, such as contractual divisions between primary and secondary care, worsening socioeconomic inequality causing mental distress, and medicolegal fears of HCPs, affected whether patients remain on APM. Themes (with illustrative quotes) are discussed below, and additional supporting quotes are available (see supplemental file 2). At the end of each theme, potential facilitators of optimal care suggested by respondents are discussed, with suggested actions for clinicians and MDPs at both local and national levels.

### 1. Confidence of HCPs around holistic management of APM

Holistic care of patients taking APMs requires HCPs trained to manage both mental and physical health concerns. Both GPs and psychiatrists lacked confidence to address both mental and physical health concerns holistically, with many patients facing fragmented care as a result, where focus was either primarily on mental health, or physical health, but rarely both. HCPs described inadequate postgraduate training and limited opportunity to retain skills after specialist qualification as contributing factors to their perceived competency gaps. Three subthemes emerged:

#### (a) GPs were reluctant to manage APM without psychiatrist support

Most GPs felt their training did not encompass APM management and therefore felt inadequately skilled to manage APM prescribing without psychiatrist support. At the same time, GPs were concerned that patients might lack confidence in their ability to advise on APMs appropriately:

> “I don’t think that my [junior doctor psychiatric] training necessarily made me more confident in relation to antipsychotics. I don’t think I’m confident enough to do that without some input.” (GP-07)

> “It’s not just a GP thing. the patient might not have the confidence in us to do it either. They’ve often demanded to be referred to psychiatry, because the GP couldn’t make them better. to be then told by the same GP ‘well actually I‘m going to stop your antipsychotic‘-the patient may not have the confidence that stopping it will be OK.” (GP-11)

Psychiatrists recognised GPs’ reluctance to manage APM when patients were discharged to primary care, but some psychiatrists felt GPs should be able to manage APM:

> “Your average GP has excellent capabilities to manage someone on antipsychotics…. The thing I think they lack… would be the confidence to do so.” (PSY-01)

Potential facilitators to help GPs manage more patients prescribed APM independently included direct support from pharmacists, who would have additional training in psychiatric medication, to support appropriate APM optimisation while remaining in primary care:

> “Psychiatry pharmacists are very easy to contact. they tend to give detailed letters as to. what you could do. There are alternatives. I wouldn‘t necessarily make a decision straight away without just sending the pharmacists a quick letter to say, “this is a situation what would your advice be” and they might say “aripiprazole and then you need to do x y z”. And that would probably give me more confidence. I don‘t think I would just switch than myself because I don‘t necessarily have the confidence or the knowledge to do that.” (GP-07)

#### (b) Psychiatrists were reluctant to address cardiometabolic risks

Respondents felt psychiatrists often disregarded cardiometabolic risks. Psychiatrists explained that they felt they lacked the necessary skills to enable them to adequately address cardiometabolic risks, but there was also a reluctance in some to consider this might be part of their role. Many respondents felt this meant a missed opportunity to intervene in cardiometabolic risk for some patients, as they acknowledged many patients with SMI might not attend for GP follow up as expected, and this puts them at greater risk of suboptimal management and poorer long-term outcomes:

> “I received no training in general practice… that‘s never been part of my psychiatric training. The idea that I would comfortably manage hypertension or diabetes… I don‘t feel comfortable with it at all.” (PSY-07)

> “It‘s all ‘go and talk to your GP‘… that‘s another step in the loop… and the patient often doesn‘t go.” (MDP-10)

GPs understood psychiatrists’ reluctance to initiate medications for cardiometabolic risks but criticised a lack of non-pharmacological interventions (e.g., referrals to dieticians or smoking cessation services), with the expectation that GPs would undertake all of these. Some also noted that psychiatrists rarely made changes to APM to ameliorate cardiometabolic risks, their focus being solely on addressing psychiatric issues:

> “There is…mention in some letters about us helping the patient control lifestyle factors… What I don‘t [see] is any evidence in letters that those referrals have been made by the psychiatric teams themselves.” (GP-11)

> “Your focus tends to be on mental health…when you‘re seeing someone in their 20s [with] acute psychotic illness and their life is coming apart - you‘re focusing on that, and you are thinking ‘they might die of heart disease when they‘re 60? If I get them to 60, I‘ve done really well!’ “(PSY-01).

Respondents felt having contracted GP support embedded in mental health teams, particularly for inpatient units to manage physical health risks, would overcome some of psychiatrists’ lack of confidence around physical health interventions, and ensure that patients had holistic care prior to discharge. Others thought non-medically qualified assistants, such as health care support workers to co-ordinate and ensure physical health monitoring occurred, would be of help in community mental health teams:

> “Either we make primary care accessible to that group of people or we embed generalists within mental health services. And I think it‘s going to be probably easier to embed people with good general physical health monitoring and physical health intervention skills within mental health, because we know that our group of patients tend to be the most resistant and wouldn‘t necessarily present to a GP”. (MDP-06)

#### (c) Gaps for HCPs in postgraduate and post-speciality training

HCPs reported inadequate training to manage both psychiatric and cardiometabolic aspects of APMs. While 14/16 (88%) GPs interviewed had undertaken postgraduate psychiatric training, no psychiatrist interviewed had any postgraduate training in general practice (Table 1). This impaired mutual understanding of each other‘s specialty: some psychiatrists thought it was a requirement that all GPs undertook psychiatric postgraduate training, and this partially drove the expectation that, therefore, GPs could manage APMs. It was of note that no psychiatrist stated they would have benefitted from postgraduate experience in primary care. Respondents confirmed postgraduate GP training did not cover APM management:

> “I think [GP trainees] were… treated differently [to psychiatric trainees]. It was felt that they didn‘t need that sort of in-depth knowledge [around APM management].” (PSY-05)

> “I would have initially thought that GPs would have all had to have done some training in [psychiatry], so am I‘m wrong in that? [interviewer explains some GPs have no psychiatric placement] … I‘m actually shocked…. I had the impression that they had to do a stint in psychiatry.” (PSY-04)

GPs were worried that other stakeholders wanted them to manage more patients on APM without appreciating the unsustainable volume of specialist secondary care work being transferred to GPs:

> “The special interest groups are saying, ‘Well, actually, this is not complicated… if you do some online teaching for an hour then the GP should know enough‘…. It‘s not possible, because we can‘t keep up to date with all the different antipsychotics… as well as keeping up to date with all the different specialities and their specialist medications.” (GP-11)

Psychiatrists confirmed they had little postgraduate specialty training on cardiometabolic risks of APM and MDPs involved in training confirmed this gap in the psychiatric curriculum. One MDP (a former director of postgraduate psychiatric training) felt that psychiatric trainees should have some exposure to working in general medicine or primary care to improve their holistic skills and gain a better understanding of how primary care functions:

> “It doesn‘t seem to be built into the curriculum. I don‘t directly influence what‘s going on in the [Royal College of Psychiatrists] anymore… I don‘t know what it says in terms of speaking to physical health…. for] adult psychiatry trainees. [we need] to give them more experience of working in. primary care, or could be on an acute medical assessment unit. I think [it] would help.” (MDP-01)

There was a concern by GPs and psychiatrists that psychiatric vocational training placements for GPs were often unsuitable to gain relevant psychiatric experience (e.g., GP trainees placed in forensic hospital posts with a low turnover of acutely unwell patients) and felt there was little recognition by both relevant Royal Colleges (who oversee postgraduate curricula for medical graduates in the UK) that management of APM was increasingly falling to GPs, as more patients were discharged to primary care. Many felt that this needed to be addressed by improved curriculum design and better co-ordination between the Royal College of General Practitioners (RCGP) and Royal College of Psychiatrists (RCPsych). Others felt there needed to be attitudinal changes within the psychiatric profession, which had to be led by the RCPsych, to accept that physical health concerns, often arising from APM side effects, was their responsibility to deal with:

*If it‘s a sequelae from their medications, I think they should be training on looking at ECGs and looking for QTc abnormalities, management of diabetes and QRISK [cardiovascular risk assessment] - I think would be sensible.* (MDP05) GPs felt continuous professional development (CPD) sessions with psychiatrists were needed, to look at holistic care of patients on APMs (with both specialities on an equal footing, rather than psychiatrists ‘lecturing GPs on what they need to do‘), reflecting the need for both specialities to develop strategies and shared protocols to improve integrated care:

> “I would love to see our [CPD] sessions return to in-person, face-to-face, group-based multidisciplinary sessions… And I would like that to be all inclusive across our health board. It‘s a brilliant opportunity that we‘ve got to develop beyond primary care.” [MDP-09]

### 2. Service pressures ‘trapping’ patients on antipsychotics in primary care

Respondents felt service pressures had increased APM prescribing, with three main subthemes emerging. Many voiced concerns about difficulty obtaining psychological interventions for patients, which meant both GPs and psychiatrists often had no recourse but to prescribe APM to help patients cope, especially for patients with non-psychotic illnesses including personality disorder, anxiety, and insomnia. Psychiatrists explained they were often under pressure to quickly discharge patients back to primary care, and this also contributed to prescribing APM to enable this to happen. However, GPs were concerned that when cardiometabolic risks were accruing in patients secondary to APM, many psychiatrists were reluctant to accept referrals back to their service for medication reviews:

#### (a) Limited psychology provision

Respondents, particularly psychiatrists, felt limited psychology provision forced APM prescribing, often used in an off-licence manner that GPs were not aware of, as a ‘stop-gap’ which was not best practice:

> “What do you do when someone is presenting as severely anxious, mood difficulties, not sleeping, incredibly chaotic: wait 18 months for therapy?… Do you simply adhere to NICE recommendations and say… ‘there‘s nothing I can do for you, you‘re going to have to wait 18 months, it doesn‘t matter how distressed you are - just deal with it?” (PSY-08)

It was felt that there needed to be more assistant psychologist posts to help manage psychological demand, due to the fact it takes 5-6 years to fully train a clinical psychologist and so few were available:

> “Some of these newer roles - the Clinical Associate in Applied Psychology, the ‘CAAPS’ roles. I don‘t want to call them the equivalent of physician associates, but they can work with people in the psychological model under supervision. They can do the psychology, the formulations, they can deliver the interventions with support and with supervision.” (MDP-01)

#### (b) Pressure to discharge patients to primary care

Psychiatrists reported pressure from service managers to discharge patients, even those with SMI if they were stable from a psychiatric perspective, and especially those who did not have a recognised psychotic illness. As a result of this transfer of patients to primary care, many GPs felt patients discharged would be unlikely to ever stop taking APM:

> “There was a lot of pressure a few years ago on caseload cleansing, and there was [a] bizarre diagram handed out: a huge pressure to [put] your caseload into one of these four boxes the only ones we should be keeping were the ‘complex and unstable‘…. and this is ridiculous if I discharged [these patients] to primary care, they‘d be back in within two weeks.” (PSY-01)

Many potential facilitators to reduce pressure to discharge patients to primary care were raised by respondents. Managers felt the removal of numerical targets imposed by governments as a measure of efficiency was needed – the preference being to measure quality of outcomes rather than numbers of patients seen - to reduce pressure on mental health teams to discharge patients. There was a recognition that many patients with chronic SMI should remain under psychiatric care, with varying degrees of input, but as a minimum, to have an annual review with a psychiatric specialist. GPs felt that proper shared-care prescribing agreements should be in place for all antipsychotic prescribing, and discharge to GPs only undertaken with mutual agreement by both services (MDP06):

> “It not really shared-care a lot of the time. So once a ‘shared-care’ is in place, secondary care often seems to think, they can then just discharge patients without any agreement… the problem is that don‘t tend to attach any funding to it. So, you don‘t end up having the ability to be able to employ a mental health nurse to do the antipsychotic reviews…I think the main difficulty is getting proper shared-care in place, with the funding was attached, and you‘ve then got the resources to do it properly.” (MDP05)

#### (c) Difficulty to get psychiatric review for patients with cardiometabolic symptoms

GPs reported difficulty obtaining psychiatric review when cardiometabolic risks developed in those patients who were discharged to their care on APM. Psychiatrists often advised ‘no change to APM’ due to pressure on their appointments, so the chance to address cardiometabolic risk (e.g., by a change of APM to one with lower propensity for side effects such as weight gain) was missed due to resource limitations, rather than a decision based on clinical need:

> “There‘s time pressure - if I‘ve got someone stable and want to switch their antipsychotic, then I‘ll have to give them more clinic appointments.” (PSY-02)

In addition to the suggestion for formal shared-care agreements for all APM use, other respondents suggested a formal mechanism for a 5-yearly specialist review of all patients taking APM long-term in primary care. There was some debate, however, as to which profession would be best placed to do these reviews, with suggestions including training GPs with a specialist interest in mental health, community psychiatric nurse prescribers, or pharmacists. Psychiatrists however, felt that it was unlikely these other professionals would have the confidence to undertake reviews to reduce or withdraw APM, so should ideally be undertaken by their own specialism:

> “For that to happen, it would need to be done by the person with the most well-developed sense of judgment of risk I think that would be a strong case to argue for it being a psychiatrist doing it. You could get somebody else doing it, but if they were less confident taking somebody off medication is always a bit of a scary thing to do. And it is always kind of tempting not to, because then you know what risks are there in doing that? So, I would suggest it might be best for a psychiatrist to do it.” (PSY-03)

### 3. Communication barriers between primary and secondary care

Respondents felt that inadequate communication, both written, verbal, and in terms of Information Technology (IT) access and integrated care, emerged as three subthemes which inhibited better integrated care of patients prescribed APMs. GPs relayed how they rarely had clear written discharge plans around ongoing APM use and struggled to get to speak to psychiatrists directly for advice. Psychiatrists related how referrals they received often lacked sufficient information to help manage patients, often from GPs who did not know the patient. All respondents, including MDPs, highlighted the fragmented plurality of different IT systems in primary and secondary care holding electronic health records (EHRs) in service silos, making it difficult for teams to access all records to get a comprehensive overview of the patient journey:

#### (a) Inadequate information in written handover

HCPs mentioned inadequate information in psychiatrist‘s letters on APM indication, duration, and monitoring; GPs were often unaware when APM was prescribed ‘off-licence’ and felt any anticipatory advice given was mostly only when to increase an APM to avoid the need to refer back to a psychiatrist. This left GPs with little confidence to make changes to APM prescribing independently:

> “As someone who completes discharge summaries, I‘m going to hold my hand up and say, we don‘t always include that… GPs get patients discharged on an antipsychotic when they‘re 23 and they‘re still on the same antipsychotic at 43” (PSY-01)

Psychiatrists reported referral letters from GPs lacked detail and lack of personal knowledge of the patient in the letter inhibited care, often only being a brief computer-collated GP summary:

> “Referral quality can be variable depending on the experience of the GP, and their interest in mental health… sometimes we [have to] go back and ask for more information.” (PSY-05)

Virtually all respondents felt that having standardised discharge and outpatient review templates that indicated not only the current treatment but clear information on indication, whether the APM was being used off-licence, intended duration, and suggested plan for switching, weaning and reduction when safe to do so, would help support GPs to make APM prescribing decisions, and that this should be performance managed as part of a shared care agreement; many felt the only focus was on initiation and dose titration, and this was insufficient to enable appropriate holistic care:

> “It would be useful if the [mental health team] said they‘re on this drug, which requires X, Y and Z monitoring, and we would expect them to stay on the drug for x length of time. And I think they should say what the indication is and that it‘s being used off-licence.” (GP10)

#### (b) Difficulties in verbal communication between primary and secondary care

GPs reported difficulties speaking to psychiatrists, with responses often from non-medical professionals unable to address their APM concerns, and psychiatrists felt GPs did not often know patients well enough to provide any useful context when they called to find out more about the patient:

> “You often find - no disrespect to those health care workers …queries that we raise are dealt with by health care workers way below the level of training of a consultant psychiatrist.” (GP-11)

> “Because of the nature of general practice, you [often cannot] talk to a GP that knows the patient personally… information that comes with the familiarity with the patient that can be difficult to access.” (PSY-07)

Some colleagues felt that services such as ‘Consultant Connect’ [27] were helpful, where a GP could speak to a senior psychiatrist for advice, but because that consultant could be located anywhere in the UK and did not know the patient or have access to the patient medical record, this was often quite limited when needing to discuss medication optimisation. Others felt that a local ‘consultant of the day’ who could take calls from GPs would be more likely to improve decision making, as they would have EHR access to guide management more effectively and document the advice given in the EHR. Psychiatrists also felt direct telephone numbers to GPs would improve communication. An indirect, but related benefit of joint CPD training was also mentioned, in that teams would get to know each other personally, which would improve mutual trust, problem-solving, and communication:

> “In [English Region], we developed a ‘psychiatrist of the day’ who would simply take calls from GP‘s, and it was amazing: the upskilling that results from that. The value of a local connection is twofold: One is, as a GP, I can seek advice about altering some of these medications, or whether or not they need referral right now.? Is there something else I can be doing with them? But two, also the ‘psychiatrist of the day’ gets to pick up what are the issues going on out there?’ What‘s the common themes that perhaps we could do something about? Do we need an education event, or a change to a pathway?” (MDP-09)

#### (c) Poor integration of IT systems used in health services

Respondents highlighted that suboptimal IT systems inhibit HCP ability to manage patients holistically, partly due to separate EHR systems commissioned by and used in primary and secondary care services. HCPs often reported inability to view EHRs, resulting in wasted time seeking information:

> “The hospital will have one electronic system, and the GP will have a second electronic system. And then psychiatry has a third, even more separate, difficult to access system, which some people have viewable access to, but not editable access.” (GP-07)

All respondents felt that improved access to shared records, utilising a common EHR system across primary and secondary care, would improve integrated care. Many would like the ability to add medications and have access to write into a common EHR, to avoid duplication of effort, and ensure that all professionals were fully up to date with patient care.

Respondents recognised that patient confidentiality needed to be protected, but some felt that an overemphasis on patient confidentiality, and having multiple EHR systems across the health service, led to fragmentation of clinical records that impaired patient care, leading to potential harm:

> “It is in every scenario - that kind of holy grail of the shared care record, it would sort it all out, it would solve it all.” (GP08)

### 4. HCPs have low expectations of patients taking APM

It became apparent that some HCPs had low expectations of patients with SMI. They found them difficult to engage with therapeutically and described patients often missing appointments, causing frustration for staff trying to help them. HCPs felt this was often due to lack of motivation in the patient group. Other HCPs, however, felt that these patients were disadvantaged by the health system, often faced discrimination, and were given insufficient information on APM use to enable informed consent. This drove some HCPs to act as advocates for their patients:

> “They are not people who ask for help…they‘re beaten down by the system as well, and they accept their lot is just s***, and ‘that‘s just the norm for me’ “They are seen to be a lost cause…. ‘Oh, they will never give up smoking, so there‘s no point in me asking‘…. And not only that, but it‘s also their own fault - it‘s the opposite of the truth, isn‘t it really…They respond well to having someone who is interested in them… I‘m not sure I make a massive difference we‘re there to pick stuff up when things go wrong, and start metformin when they inevitably become diabetic, or write the death certificate when they die young. You know, it feels a bit depressing to just verbalize it.” (GP-08)

Many HCPs felt that tackling lowered expectations towards patients with SMI was challenging, and this mirrored societal attitudes. However, one MDP who represented patients from a mental health charity, did report that they felt GPs who had undertaken psychiatric training were more likely to be able to challenge and act as advocates for patients, and that postgraduate psychiatric training should be mandated in the GP training programme:

> “I think with GP training, you maybe get one opportunity to spend time on a mental health ward, and I know from speaking to GPs, that those GP‘s that have done that understand the system better in terms of referrals… they are better able to challenge it and to be an advocate on behalf of their patients.” (MDP02)

### 5. Strategic factors affecting APM prescribing in primary care

Participants identified multiple professional and organisational constraints acting as barriers to optimise APMs. Three main subthemes emerged; the first subtheme focussed on the difficulties with the contractual split between primary and secondary care, with the UK GP General Medical Services (GMS) contract and narrow criteria for patient inclusion on the Quality and Outcomes Framework (QOF) mental health register, meant many patients now prescribed APM in primary care for non-psychotic illness were not being monitored effectively. Organisational competition and ‘protectionism’ for resources were mentioned, meaning patients could be disadvantaged by siloed care largely dictated by organisational budgetary constraints. A second theme centred around socioeconomic deprivation and medicalisation of distress driving APM use. A final theme described the fear many HCPs had around stopping or changing APMs in case an adverse event occurred that would render them liable to ‘societal blame’ or censure by regulatory bodies:

#### (a) Contractual divisions between primary and secondary care

Psychiatrists and MDPs felt the UK General Medical Services (GMS) GP contract structure inhibited shared care:

> “Sometimes it‘s the GMS contractual issues… that constant argument around “I‘m not going to do this unless [GPs] get paid for it”. Those sorts of issues really need to be addressed… sometimes we forget there‘s a patient at the end of all these decisions.” (MDP-08)

The Quality and Outcomes Framework (QOF) in the UK GMS contract incentivises GPs to monitor physical health risks for patients with SMI, but incentives have been largely abolished in GMS contract revisions in Wales and Scotland; GPs felt this would deter ongoing monitoring. Many were concerned the QOF ‘mental health register’ criteria, being focussed almost exclusively on psychotic illness such as schizophrenia or bipolar affective disorder, did not capture all patients prescribed APM, so there was a risk many patients were not reviewed comprehensively:

> “In England, we still have the [QOF] mental health register. but it doesn‘t pick up people who‘ve got a personality disorder.” (GP-04)

Respondents reported tensions between organisations contributed to protectionism that impaired care. A former Royal College officer reported conflicts of interest between representing members’ interests, and serving patients:

> “There is a real tension between being a member‘s organization that needs to deliver for its members’ interests, and doing the right thing…, I was there representing [Royal College] we can see what the right thing is to do, but as membership organizations, we have to protect the needs of our members. So, you would not expect the Royal College of GPs to meet with the Royal College of Psychiatrists and [say] “You have all the money we get for mental health, and you just sort out everything”…protectionism gets in the way of better care.” (MDP-04)

Some respondent*s* felt influence of patient groups and political targets could skew resource allocation:

> “When… you‘re taking a massive stack of antipsychotics, with all the negative impact that has on motivation. those patients are the ones that I feel are most neglected by mental health services the patient groups that talk the loudest, are [those] who we perhaps have the least value to offer. the patient groups who have the smallest voice. need the most attention.” (PSY-07)

MDPs felt frustrated attempts to address APM overprescribing did not result in savings to reinvest in services such as psychology:

> “Prescribing budgets are held in a different part of the [NHS organisation] … I can‘t get hold of prescribing budgets, even if I‘ve saved on one element of them to reinvest the savings.” (MDP-06)

Many respondents felt that contractual divisions between primary and secondary care created barriers, for example around ability to refer to weight management clinics being restricted to primary care, and service commissioners and providers needed to facilitate pathways that allowed ease of referral for interventions from all HCPs. Others felt that there was a need for more joint working between Royal Colleges, the British Medical Association and service providers to identify gaps in holistic provision and the need to alter the funding mechanisms to overcome these. Many HCPs highlighted that APM prescribing should indicate ongoing monitoring, rather than specific SMI diagnoses, as currently occurred in the QOF mental health register:

> “I think the Royal colleges and others come together really well on single pieces of work, but actually our chances of getting the Academy of Royal Colleges to say, “Primary care is the way forward. Disinvest from all your fancy hospital services and spend the money where they make the biggest difference”, there is zero chance, even though every leader in the Academy of Royal Colleges can intellectually appreciate the benefit.” (MDP04)

#### (b) Worsening inequality driving antipsychotic use

Respondents often felt antipsychotic use was a ‘stop-gap’ to deal with consequences of socioeconomic inequality and the medicalisation of distress, which health services could not address directly, and the patient primarily needed social interventions regarding finances, housing, or education:

> “We go at stuff from the health level view. Until we‘re able to deal with the fact [people] can‘t feed their kids, or heat their homes, that schools don‘t have the resources to put into troubled young people. [antipsychotic prescribing] is not going to reduce in our medicalized society. I can absolutely see why [antipsychotic prescribing] rates are rising.” (MDP-04)

Another common area of concern was around pressure from relatives and care home staff to prescribe APMs to manage challenging behaviours in older adults with dementia, and felt this was largely down to inadequate training, retention and recruitment of carer staff who have the skills to manage patients with behavioural difficulties using non-pharmacological methods. Many respondents felt powerless to address the wider socioeconomic determinants driving increasing mental distress in the community but recognised that this had to be addressed as a political priority that encompassed services beyond health. A number however felt that investment in better training and retention of skilled care home staff would reduce APM prescribing in elderly patients:

> “So, for example, care homes are running on empty. They don‘t have people who are skilled…. they have token specialist dementia champions, but they just don‘t have the resources to sit with people while their anxiety levels reduce, or engage them with memory rooms, and things that we know improve or reduce the need for antipsychotic use.” (GP-04)

#### (c) Medicolegal and regulatory fears of HCPs

HCPs mentioned fears around stopping APMs if a patient subsequently harmed themselves or others; some felt they ‘prescribe to be seen to do something’ due to working in an unforgiving regulatory culture:

> “If I was to stop an antipsychotic…if something adverse happened, I think I would have a very difficult time justifying that action, even if it‘s a relatively safe thing to [do]… those things lead to medication overprescribing… even if it‘s the wrong thing to do, it‘s almost a sense that having done something is better than doing nothing.” (GP-04)

Psychiatrists reported unrealistic societal expectations that an adverse outcome must ‘always be someone‘s fault’ which inhibited their willingness to deprescribe:

> “Physicians accept that there‘s a certain percentage of COPD patients who will die, that it doesn‘t matter much resource you use…they will get worse, and some of those people will die. And then there is a very marked reluctance… [within society] to accept that [psychiatric] conditions also have a morbidity and mortality… which I find frustrating.” (PSY-07)

Many respondents felt that, at times, a degree of over-regulation created barriers, in that it inhibited ability to devise appropriate care pathways and encouraged defensive clinical practice. They mentioned that the regulators did not seem to recognise that this stifled ability to innovate at times and did not seem to recognise that overzealous regulation could paradoxically lead to indirect patient harm, and so wanted a debate around the culture in which they had to practice.

One final unintended consequence of overzealous performance management that appeared to contribute to increased APM use was around prescribing indicators for benzodiazepines being closely monitored by national medicines management teams, with targets being to achieve a reduction in their use. As a result, this was often given as feedback to clinicians in quarterly reports on their prescribing activity, and many felt unfairly scrutinised as a result. Some practitioners thus ‘gamed they system’ by switching to using sedating APMs (e.g., quetiapine) to treat insomnia rather than using benzodiazepines, as this was not monitored at the national level, and thus some MDPs suggested that the wider use of off-licence APM needed to be monitored also:

> “I‘m guessing if we use diazepam as an example - diazepam was the elixir for everything. And we had, going back, a massive uptake in the prescription of diazepam. We are aware of the issue of diazepam now has, in a sense of its addiction… so we just turn to other medication with a similar effect; in this case it is antipsychotics.” (PSY-04)

## DISCUSSION

### Summary

Our research shows how HCPs and MDPs reported competency gaps that impaired HCP ability to manage the varied mental and physical health needs of patients’ prescribed APMs appropriately. Stakeholders suggested that deficiencies in postgraduate experience and professional development contributed to GPs and psychiatrists’ lack of confidence to manage both the mental health and physical health aspects of APM management. Lack of postgraduate GP experience for psychiatrists inhibited their understanding of GP competences. Communication barriers, lack of integrated care records, limited resources, particularly in relation to psychology provision, and perceived pressures to move patient care back to GPs ultimately resulted in patients in primary care being ‘trapped’ on APM, inhibiting opportunities to deprescribe, and increasing cardiometabolic risk in the longer term. Organisational and contractual barriers between services exacerbated risks, with socioeconomic deprivation driving overprescribing. Both GPs and psychiatrists voiced professional fears of being blamed if an event causing harm occurred after stopping an APM, which inhibited deprescribing confidence. Many respondents had low expectations of patients with SMI, which also contributed to care deficiencies. These factors act at several points in the care pathway and contribute to the increasing prevalence of APM use and accretion of cardiometabolic risk in this patient group in primary care (Figure 1).

**Figure 1:**
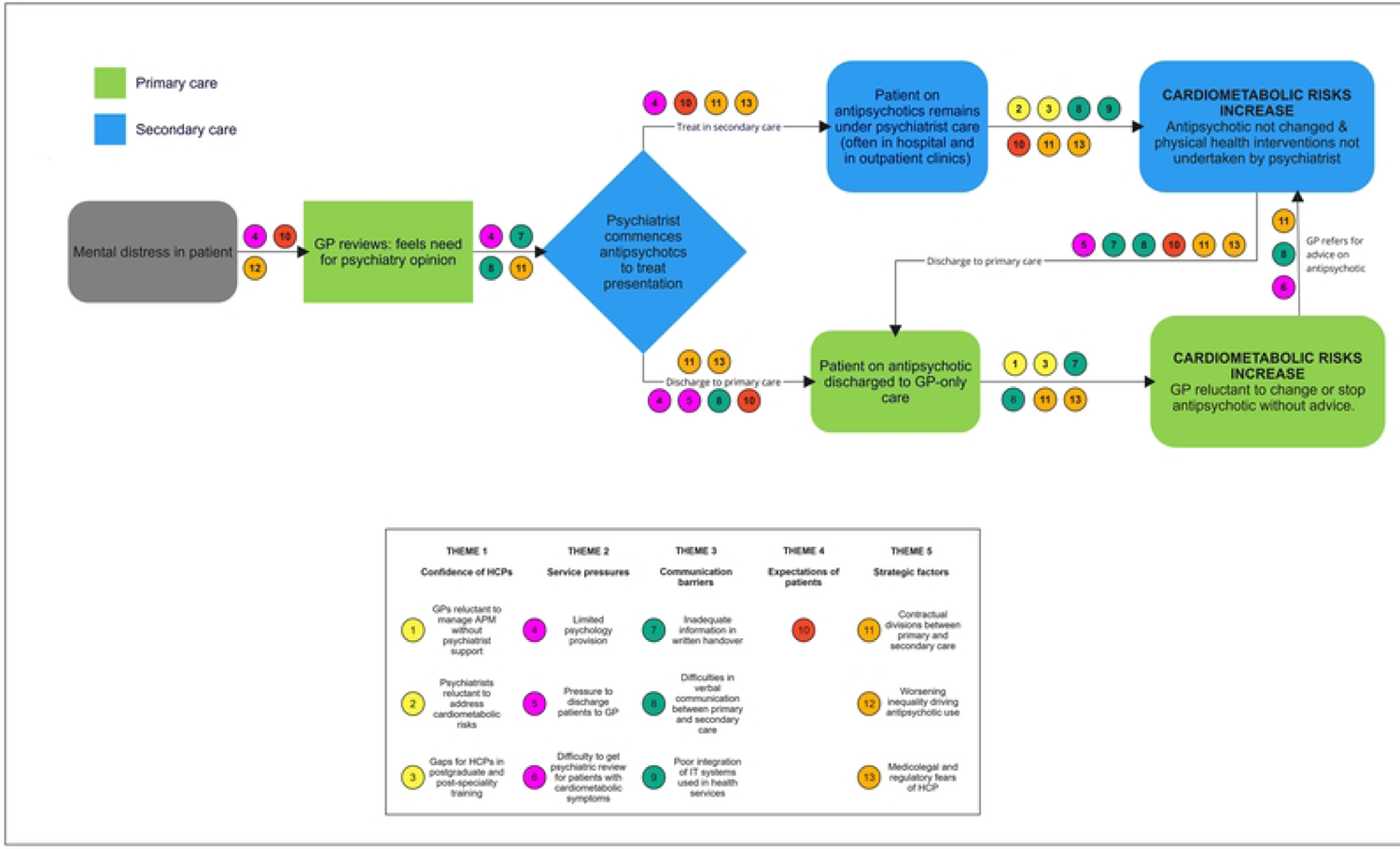
Pathway of patient care on antipsychotics and barriers to better integrated care

### Strengths and Limitations

This study examines reasons for increasing APM prevalence in primary care. We had input from team members from different professions (medicine, pharmacy, health research), medical specialities (general practice, psychiatry, clinical pharmacology, public health) and the study sought views of those managing services, helping shape mental health policy and contractual arrangements, and involved in postgraduate training. We recruited participants from across three UK nations, to minimise bias that might result from one health region.

Data saturation was achieved. However, participation, which was voluntary, may mean participants had a particular interest in the problem, and we may not have acquired views of less interested clinicians, so there is a risk of recruitment bias. Finally, this is a qualitative study that identifies perceived issues of concern in relation to APM management, but it does not provide information about the relative importance of the different themes identified. Also, the work is undertaken in the UK, and while there may be similarities with other high-income countries there will also be differences worthy of exploration to better understand common international issues. Finally, this study does not seek the views of patients who are prescribed APM and managed solely in primary care; a separate qualitative study examining this is currently underway.

### Comparison with existing literature

This is the first study to examine management of APM for patients discharged to primary care, which makes comparisons difficult, but other studies partially consider this. Nash *et al* [20] examined decision-making around APM switching by psychiatrists and GPs in one health region in England but did not specifically consider patients managed solely in primary care. This work and other studies conducted with patients and professionals in the Netherlands reported similar themes: concerns around jeopardising mental health by switching APMs to reduce cardiometabolic risk was mentioned as a reason not to do so, poor engagement with patients to make informed decisions, and diffusion of responsibility with respect to managing cardiometabolic risks secondary to APM use [5, 20]. Several studies in the UK, Netherlands and Canada confirm difficulties in communication and collaboration between primary and secondary care inhibits integrated care for people taking APM [20, 28–31]. Moncrieff *et al* examined patient and HCP factors acting as barriers to deprescribing of APM in the UK [32] and reported concerns about relapse and potential harms to the patient, but this did not elucidate concerns of the HCPs around medicolegal or regulatory risks to their professional role. Our data confirms these findings, and extend them, showing that factors including HCP training, contractual, organisational, and medicolegal factors also act as barriers to better person-centred care in the UK.

### Implications for future research and clinical practice

Our findings suggest several practical actions policymakers and practitioners can take now to improve person-centred care for people on APM, which we summarise in Figure 2:

**Figure 2:** Potential actions to improve integrated care for patients taking antipsychotics discharged to primary care

This will require action at several organisational levels, including changes to postgraduate curriculum design for both general practice and psychiatry, evidence-based guidance on APM prescribing duration in non-psychotic illness, greater investment in non-pharmacological resources and input from professional regulators to address HCP fears around a ‘blame culture’ that exists currently. Our analysis suggests that addressing competency gaps and communication barriers between primary and secondary care are those which are likely most amenable to change locally within health service organisations. Implementing structured discharge letters and mandatory use of shared care protocols are relatively easy to implement to improve integrated care of patients taking APM. Psychiatric inpatients usually have considerable physical health data collated during admission that allows accurate quantification of cardiovascular disease risk, but these are not always acted on by psychiatric teams; training to ensure appropriate pharmacological interventions are undertaken is needed [33]. Modification of care pathways for non-pharmacological interventions to allow referral by mental health staff, regular reviews by competent HCPs able to optimise or withdraw APM, and appointment of staff to provide support to patients that is non-pharmacological may be harder to achieve. Particular challenges exist in terms of capacity in terms of psychological provision, but also within in the social care sector with staff capable of managing behavioural consequences of dementia without recourse to APM use. The wider societal factors, around attitudes towards patients with SMI and dealing with socioeconomic factors increasing mental distress are beyond local action and require a co-ordinated national approach. It is also clear that well-intentioned performance management and regulation may paradoxically contribute to increasing prevalence of patients on APM in primary care. HCPs increasingly use APM as an alternative to benzodiazepine use for insomnia and anxiety, due to the latter being closely monitored as a quality prescribing indicator in some regions; medicines management teams should consider the wider use of APM for non-psychotic illness in their performance indicators. The wider regulatory milieu in which HCPs perceive they are working is leading to defensive practice, where patients are left on APM due to fear of professional censure should a catastrophic adverse event occur if HCPs reduce or withdraw the medication. Regulators need to reassure HCPs that this is unlikely to occur, as this was a common concern for HCPs; a national debate on this is required.

Further research is required to explore experiences of patients prescribed APMs discharged to primary care, and to examine if similar barriers to person-centred, integrated care exist internationally in other health systems where primary and secondary care operate in an analogous manner to the UK (e.g., Australia, New Zealand, Ireland, Canada). Studies have examined attitudes of patients taking antipsychotics regarding their management and physical health [30], but none formally examined the growing proportion of patients prescribed APM no longer under psychiatric review. Further study is needed of clinical and postgraduate educational policies in the UK and elsewhere that have contributed to fragmented services with HCPs, particularly in secondary care having inadequate understanding of the primary care context. The theme of regulatory fear is also evident in HCPs treating patients with SMI and acts as a barrier to deprescribing of APM. This needs further exploration internationally, with regulators, patients, and HCPs together – to consider how the regulatory and performance management ‘observer effect’ may impair or distort clinical practice.

## CONCLUSIONS

There is considerable risk that the increasing use of APM in society is inadequately managed. Unless we address the factors that contribute to fragmented and suboptimal care for those taking APM, poor long-term outcomes in terms of increased rates of cardiometabolic disease will be difficult to ameliorate. The theme of regulatory fear by HCPs is a new factor emerging as a barrier to deprescribing of APM, and this needs further examination in other studies to determine if this is an international concern. Many changes could easily be made to improve person-centred, integrated care of those on APM with minimal need for change by health service organisations. However, other issues such as inadequate availability of non-pharmacological alternatives to APM and socioeconomic factors increasing mental distress will require national leadership by key stakeholders to improve the current situation.

## Data Availability

All relevant data are within the manuscript and its Supporting Information files.

## Acknowledgements

We thank the generous input of the participants who took part in the interviews.

## Contributors

AW, FM, SS, IB and LW conceived the study and wrote the application for funding and the protocol. PS provided public perspective on the study design and results. AW conducted the data collection. AW, AA, SW and LW undertook the primary coding and thematic analysis, and all other authors (PA, FM, SS, DJ, IB) contributed to the data analysis and discussion. AW and LW drafted the manuscript, and all authors commented on the manuscript.

## Consent for Publication

Not Required.

## Provenance and Peer Review

Not commissioned; externally peer reviewed.

## Supplemental material

Study protocol, semi structured interviews template, additional quotes.

## Open access

Put in relevant licence.

---

| Theme | Sub-theme | Problem | Action possible now? | Suggested solution | Action to achieve by: |
| --- | --- | --- | --- | --- | --- |
| Confidence of HCPs around holistic management of APM | GPs reluctant to manage APM without psychiatrist support | GPs Lack confidence to trial APM optimisation | Yes | Structured letters* from psychiatrists give clear advice on indication, duration, switching, and dose of APMs before discharge, and confirm discussion with patient. | Directors of Mental Health services |
|  | Psychiatrists reluctant to address cardiometabolic risk | Psychiatrists not addressing physical health risk | Yes | Support for psychiatrists: Employment of GPs in advisory roles to support cardiometabolic interventions while under psychiatric care.<br>Support staff to co-ordinate physical health monitoring and interventions | Directors of Mental Health services |
|  | Gaps for HCP in postgraduate and post-speciality training | Training does not cover the full range of skills to provide holistic care | No | Co-ordination of postgraduate curriculum design by Royal College of General Practitioners and Royal College of Psychiatrists<br>Mandatory primary care experience in postgraduate training for all psychiatry trainees. | National priority setting by Royal Colleges |
| Service pressures 'trapping' patients on antipsychotics in primary care | Limited psychology provision | Lack of psychology driving APM use | No | Employ clinical associates in applied psychology to increase psychological therapy capacity.<br>Savings in psychotropic medication use earmarked for investment in psychological provision. | Accountable officer of health providers;<br>Directors of Mental Health services |
|  | Pressure to discharge patients to primary care | Limited psychiatric appointment resource | No | Formal shared care agreements for antipsychotics before a patient can be discharged to primary care with mandated plan on future cessation and withdrawal. | National priority setting |
|  | Difficulty to get psychiatric review for patients with cardiometabolic risks | Limited psychiatric appointment resource | No | Formal shared care agreements to include a 5 yearly mandated review by psychiatrist to determine if ongoing antipsychotic use is needed.<br>Train pharmacists to proactively address cardiometabolic risk in pharmacy and GP settings | National priority setting |
| Communication barriers between primary and secondary care | Inadequate information in written handover | Inadequate advice in discharge letters to GPs | Yes | Structured letters* (see above). | Directors of Mental Health services |
|  | Difficulties in verbal communication between primary and secondary care | Barriers to verbal communication | Partly | Direct telephone lines to "duty medical professional"; use of alternative communication (e.g., Microsoft Teams) to make contact easier. | Directors of IT, Mental Health services, Primary Care |
|  | Poor integration of IT systems used in health services | Inadequate access to care records | Partly | Ensure all clinicians have a minimum of 'read access' to summary care records (GP and mental health records)<br>Adoption of a common electronic health record system across the primary and secondary care estate with shared read-write access. | Directors of IT |
| HCPs have low expectations of patients taking APM | Low expectations included feeling patients won't engage, are not motivated, and discriminatory risks | Attitudes of HCPs | Partly | Challenge attitudes and look at different ways to engage patients with SMI which reduce inequality of access to care | Directors of mental health services and primary care;<br>National priority setting |
| Strategic factors affecting APM prescribing in primary care | Contractual divisions between primary and secondary care | QOF registers inadequate | Partly | Ensure each GP practice has register of patients on antipsychotics with performance management of monitoring and use by commissioners of services | Directors of primary care; National priority setting |
|  | Worsening inequality driving antipsychotic use | Socioeconomic deprivation causing distress | No | Redistribution of resources to improve mental health resilience. | National priority setting |
|  | Medicolegal and regulator fears by HCPs. | HCP fear of censure | Partly | Regulators to reassure healthcare professionals; set up a joint committee to investigate and report on unintended harms of the regulatory environment impacting on psychiatric patient care.<br>Consider use of antipsychotics in non-SMI illness for prescribing performance indicators. | National priority setting;<br>Medicines management teams |

